# Increased Neonatal Deaths in Texas After SB8, a Cardiac Activity-Based Abortion Ban

**DOI:** 10.1101/2024.06.26.24309400

**Authors:** Jeremy Samuel Faust, Ji Chen, Benjamin Renton, Onyekachi Otugo, Alexander Junxiang Chen, Miranda Yaver, Zhenqiu Lin, Sonja A. Rasmussen, Harlan M. Krumholz

**Affiliations:** Department of Emergency Medicine, Mass General Brigham; Division of Health Services Research, Harvard Medical School, Boston, MA; Yale New Haven Hospital, Center for Outcomes Research and Evaluation, New Haven, CT; Ontos Analytics, Cambridge, MA; Harvard University, Cambridge, MA; Department of Political Science, Wheaton College, Norton, MA; Department of Genetic Medicine, The Johns Hopkins University School of Medicine, Baltimore, MD; Yale University, Center for Outcomes Research and Evaluation, New Haven, CT

## Abstract

**Introduction:** On September 1, 2021, Texas enacted Senate Bill 8 (SB8), a law banning abortions after the detection of fetal heart activity, without exceptions for early detectable fetal abnormalities with low survival probability. The effect on neonatal mortality is unknown.

**Methods:** We deployed difference-in-differences (primary analysis), and Joinpoint and seasonal autoregressive moving integrated averages (sensitivity analyses) to determine whether all-cause mortality among Texas neonates increased after SB8 was enacted, and compared the findings to states with permissive abortion laws. Changes in mortality due to congenital malformations, deformations and chromosomal abnormalities were also measured.

**Results:** Compared with states where abortion remained legal, approximately 177 all-cause excess neonatal deaths (2.3 per 10,000 births, interaction p<0.003) occurred in Texas. A sensitivity analysis using Joinpoint logistic regression spontaneously identified a mortality trend change in Texas coinciding with SB8 enactment. Approximately 113 excess deaths (1.5 per 10,000 births, interaction p<0.001) due to congenital malformations, deformations and chromosomal abnormalities occurred among Texas neonates after SB8 enactment. Compared to expected deaths based on prior internal trends (sARIMA), Texas recorded 289 (95% CI 18-559) excess neonatal deaths (3.7 per 10,000 births; 95% CI 0.2-7.2) in 2022-2023. A preliminary analysis of post-SB8 shares of neonates listed as “not alive” on birth certificates by race/ethnicity found increased relative contributions from non-Hispanic White and Hispanic populations.

**Conclusion:** Texas SB8 was responsible for between 177-289 excess neonatal deaths from Quarter 4, 2021-Quarter 4, 2023. These results suggest that if other SB8-like abortion bans are enacted, increases in neonatal mortality may occur elsewhere.

## Introduction

On September 1, 2021, Texas enacted Senate Bill 8 (SB8), a law banning abortions after the detection of fetal heart activity, without exceptions for early detectable fetal abnormalities with low survival probability.^1^ We hypothesized that mortality in Texas neonates (ages 0-27 days) might increase, given increased driving distances to the nearest legal abortion facility after SB8 enactment and further after the *Dobbs v. Jackson Women’s Health* decision.^2^

## Methods

Accordingly, we deployed difference-in-differences analyses comparing neonatal mortality trends in Texas to states where abortion remained legal (Table S1). Separately, we used seasonal autoregressive integrated moving averages (sARIMA) to measure excess deaths by comparing observed to modeled expected deaths by jurisdiction (i.e., internal synthetic controls) (See Supplementary Methods).

## Results

After SB8 (Quarter 4, 2021-Quarter 4, 2023), a long-standing downward trend in neonatal mortality in Texas reversed (Figure, Panel A). Compared with states where abortion remained legal, approximately 177 all-cause excess neonatal deaths (2.3 per 10,000 births, interaction p<0.003) occurred in Texas. A sensitivity analysis using Joinpoint logistic regression spontaneously identified a mortality trend change in Texas coinciding with SB8 enactment (Supplementary Methods, Figure S1, Table S2). Approximately 113 excess deaths (1.5 per 10,000 births, interaction p<0.001) due to congenital malformations, deformations and chromosomal abnormalities occurred among Texas neonates after SB8 enactment (Figure, Panel B). Compared to expected deaths based on prior internal trends (sARIMA), Texas recorded 289 (95% CI 18-559) excess neonatal deaths (3.7 per 10,000 births; 95% CI 0.2-7.2) in 2022-2023 (Table S3). A preliminary analysis of post-SB8 shares of neonates listed as “not alive” on birth certificates by race/ethnicity found increased relative contributions from non-Hispanic White and Hispanic populations (Figure S2).

**Figure.**
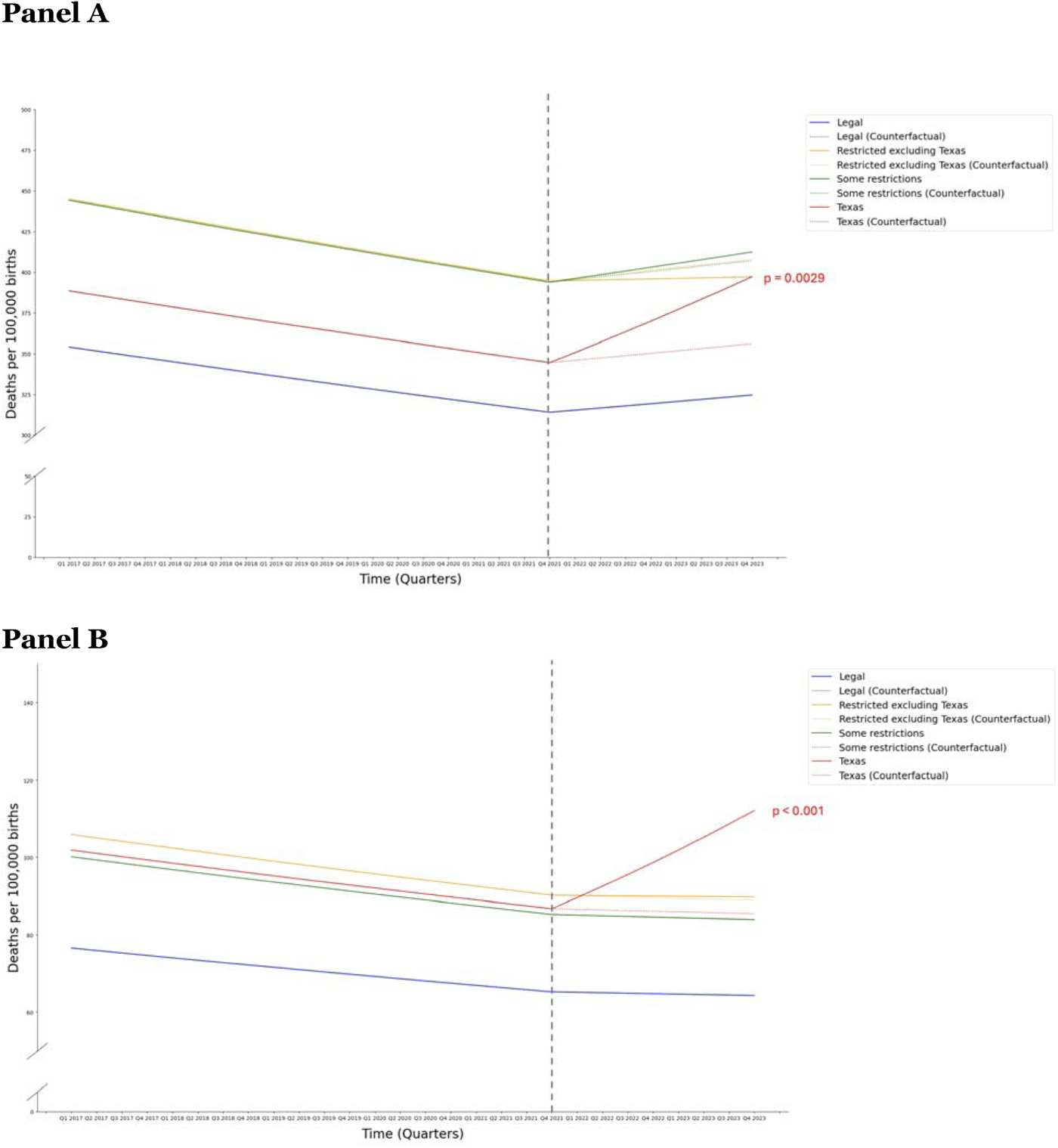
Trends in neonatal deaths by difference-in-difference logistic regression. **Panel A:** Fitted all-cause deaths per 100,000 births in Texas (red line), Restricted (excluding Texas) states (yellow line), Some restrictions states (green line), and Legal states (blue line) are shown. Counterfactual “expected” deaths for Texas, Restricted (excluding Texas) states, and Some restriction states are shown as dotted lines for their corresponding jurisdiction, and are based on the post-SB8 fitted mortality slope observed in Legal states. Interaction p values were significant for Texas compared to Legal states (P=0.003), but not for Restricted (excluding Texas) states (p=0.380), or Some restrictions states (p=0.642). **Panel B:** Fitted deaths due to congenital abnormalities (*International Diseases Classification-10* Chapter Q00-99) per 100,000 births. Interaction p values were significant for Texas compared to Legal states (P<0.001), but not for Restricted (excluding Texas) states (p=0.886), or Some restrictions states (p=0.983).

## Discussion

The difference in SB8-associated mortality across the difference-in-differences and sARIMA analyses likely reflects complementary comparator strengths. The former used observed contemporaneous mortality in states where abortion remained legal; the latter used Texas trends and modeled expected deaths had there been no change with SB8. Difference-in-differences approaches permit causal inference while synthetic internal control excess mortality analyses (e.g., sARIMA) can minimize spillover effects, in which policies in one jurisdiction effects behaviors in others.

Contrasting previous reports assessing abortion restrictions by race/ethnicity, preliminary analysis found increased relative contributions to neonatal mortality from non-Hispanic White mothers (among whom most abortions ≥21 weeks of gestation occur).^3,4^ This might reflect loss of abortion access regarding fetuses with early detectable fatal congenital abnormalities.

Higher neonatal mortality has previously been reported in states with restrictive abortion policies.^5^ These results suggest that if other SB8-like abortion bans are enacted, increases in neonatal mortality may occur elsewhere.

## Data Availability

All data produced in the present study are available upon reasonable request to the authors

## Acknowledgements

The authors wish to thank Kristina Fiore for suggesting the study.

