## Supplemental Information for "Increased Neonatal Deaths in Texas After SB8, a Cardiac Activity-Based Abortion Ban"

**Supplemental Methods.**

Natality data were acquired from CDC Natality WONDER.^1^ Mortality data for the difference-in-differences analyses and sARIMA excess mortality modules were acquired from CDC WONDER with the exception of the race/ethnicity analysis (see below for details, page 2).^2,3^

**Difference in Difference analysis.**

We estimated the association between the enactment of SB8 in Texas and mortality by conducting a difference-in-differences analysis of pre- and post-policy mortality slopes in comparison to other state groupings.^4^ For state grouping rationales, see Table S1. Analyses were conducted for all-cause neonatal mortality and for deaths caused by congenital malformations, deformations and chromosomal abnormalities (*International Classification of Diseases-10* Chapter Q00-Q99). Specifically, we assume that the mortality rate $p_{ij}$ for state group $i$ at time $j$ follows a logistic regression, and the model incorporates time (quarterly), state group, a time-varying variable indicating the time since the policy enacted, and the interaction term of each state group and the time-varying variable; that is,

$$\mathrm{logit}\left( p_{ij} \right)=C_{ij}+\alpha_{0}+\alpha_{1}t_{ij}+\alpha_{2}\mathrm{StateGroup}_{i}+\beta\left( t_{ij}-t_{SB8} \right)_{+}+\gamma\cdot\mathrm{StateGrou}p_{i}*\left( t_{ij}-t_{SB8} \right)_{+},$$

and $t_{+}=\max\left\{ t,0 \right\}$, $C_{i}$ is a known offset term for State group $i$. In the model, $\left( t_{ij}-t_{SB8} \right)_{+}$ is to evaluate the change in mortality rate slope after enactment of the policy; the interaction term $\mathrm{StateGrou}p_{i}*\left( t_{ij}-t_{SB8} \right)_{+}$ is to evaluate whether the post-policy change in mortality rate slope for each state group differs from the reference group (Legal States group). The difference is considered significant if the interaction p-value < 0.05.

After model fitting, the observed mortality rate and counterfactual mortality rate (assuming the trend in each state group would be the same as that in the reference group during the post-policy period) are estimated for each quarter. The excess mortality rate for each quarter is then calculated as the observed rate minus the counterfactual rate. Subsequently, for each state group, the total number of excess deaths and the overall excess mortality rate in the post-policy period are calculated, taking into account the number of births.

**Joinpoint analysis.**

To assess the robustness of our primary difference in difference analysis, joinpoint analysis was performed as a sensitivity analysis.^5^ This approach aimed to determine whether a joinpoint would be spontaneously identified at the time relevant to SB8 policy enactment. In the joinpoint analysis, the joinpoints $\tau_{1}<\ldots<\tau_{K}$ are assumed to be unknown and detected by the model automatically using stepwise model selection techniques. The data for each state group is fitted separately by the following model:

$$\mathrm{logit}\left( p_{j} \right)=C_{j}+\alpha_{0}+\alpha_{1}t_{j}+\sum_{k=1}^{K} \beta_{k}\left( t_{j}-\tau_{k} \right)_{+} .$$

A significant joinpoint detected from the model indicates that the mortality rate slope has changed significantly at an identified joinpoint. As the model is fitted for each state group separately, to control the family-wise error rate, Bonferroni correction is applied to adjust the p-value. The joinpoint is significant if the corresponding adjusted p-value < 0.05.

**All-cause and cause-specific excess mortality by seasonal autoregressive integrated moving averages (sARIMA) analysis.**

All-cause excess mortality in the neonatal population (ages 0-27 days) was defined as raw observed deaths minus modeled expected deaths divided by number of births. To calculate expected mortality, we trained seasonal, autoregressive integrated moving averages (ARIMA) models on five full pre-SB8 years of quarterly death counts (January 1, 2017-December 31, 2021), using population (natality) as a covariate to overcome stationarity (thus account for the fact that expected deaths are dynamically influenced by the number of births). Analyses were carried out for each of the jurisdictions assessed in the difference-in-differences module. Therefore, for each jurisdiction, expected deaths were determined by comparison to the counterfactual for that jurisdiction (i.e., individual internal synthetic controls). Monthly excess mortality (to enable capture of seasonal trends) was determined, batched into quarters, and subsequently summed for individual years (2022 and 2023), and for the total post-intervention study period (2022-2023). Model selection was determined using Akaike information criterion. 95% confidence interval boundaries were derived directly from the sARIMA model using the auto.arima function in the R statistical software. The cumulative period CI boundaries were obtained through the 5,000 simulation samples from the estimated sARIMA model for quarters, each year (2022 and 2023), and the total study period (2022-2023). Note: in contrast to the difference-in-differences analysis, in this assessment, we used January 1, 2022, as the start of the post-SB8 period. This was done to account for the time between when fetal abnormalities with low survival probability would normally be detected during pre-natal care (pregnancies in the second trimester when SB8 was enacted) and when those pregnancies would reach full term (early 2022), similar to the rationale described by Bell *et al*, (JAMA. 2023;330(3):281-282. doi:10.1001/jama.2023.12034).^6^ An *ad hoc* sensitivity analysis of the difference in difference module using January 1, 2022, as the start date did not change the overall findings.

**Race/ethnicity analysis.**

An analysis of changes in newborn mortality by race/ethnicity was conducted using CDC Natality WONDER. (CDC WONDER for mortality does not permit queries that combine race/ethnicity information and infant age groupings limited to 0-27 days.) However, Natality WONDER provides yes/no outputs for “Infant Living at Time of Report” data at the time of the birth certificate report. Demographic information regarding these infants be queried by the reported race/ethnicity of the father, mother, or both. We selected reported maternal race/ethnicity. We then determined the contributed shares by race/ethnicity of infants listed as not alive (i.e., infant not alive at time of report) on birth certificates before (2017-2021) and after (2022-2023) the enactment of SB8 in Texas. Adequate data were available for non-Hispanic Asian, non-Hispanic Black or African American, Hispanic, non-Hispanic White demographics. Pearson’s chi-square testing was deployed to determine whether the shares of infants not alive at the time of birth certificate changed after SB8 enactment.

**Table S1. State grouping rationales.**

| State | Ban Status | Confirmed | Rape/Incest Exception | Notes |
| --- | --- | --- | --- | --- |
| Alabama | Ban | Yes | No | Alabama Human Life Protection Act went into effect June 2022, only limited exceptions to save the woman’s life |
| Arkansas | Ban | Yes | No | Arkansas AG announced on 6/24/22 that its trigger law was enacted; 0 legal abortions performed post-Dobbs |
| Georgia | Ban | Yes | Yes | 6-week abortion ban went into effect right after Dobbs; in 11/22, a district court struck down its key provisions as unconstitutional, but 1 week later it was reinstated pending appeal |
| Louisiana | Ban | Yes | No | Near-total ban signed into law in 6/22, on 6/27/22 a judge blocked enforcement issuing a TRO, but it went back into effect on 7/8; new TRO issued on 7/12/22 but on 7/29/22 a judge reinstated the ban pending litigation, in 8/22 the LA Supreme Court rejected appeal, leaving the ban in place |
| Mississippi | Ban | Yes | Yes | Near total abortion ban (trigger law) went into effect 7/7/22. Judge declined to block the law from going into effect. |
| Missouri | Ban | Yes | No | Missouri the first state to pass near-total trigger ban following Dobbs (minutes following decision) |
| North Dakota | Ban | Yes | Only up to 6 weeks | Trigger ban signed into law but challenged in court, then state’s only abortion provider moved from ND to MN; state district judge temporarily blocked the law in 7/2022, and in 3/23 a court upheld the decision; Gov signed near total ban in 4/23 |
| Oklahoma | Ban | Yes | No | Trigger law (near total) went into effect immediately post-Dobbs + 1910 abortion ban in effect post-Dobbs; OK Supreme Court struck down in 5/23 two of OK’s abortion bans passed in 2022 but because of the 1910 law abortion remained illegal in OK |
| South Dakota | Ban | Yes | No | Abortion ban at any point in pregnancy passed in 2006 went into effect following Dobbs |
| Tennessee | Ban | Yes | No | Heartbeat law in effect in 6/28/2022; Trigger ban with just exception for life of woman went into effect 8/25/22, new law in 4/23 carving out narrow exceptions; all abortion clinics closed or ceased abortion services in TN |
| Texas | Ban | Yes | No | After a TRO, the TX Supreme Court ruled in early July 2022 that Texas could enforce its 1925 abortion ban; Trigger near-total ban went into effect 8/25/22; SB8 also in effect |
| Wisconsin | Ban | Yes | No | Long-dormant 1849 abortion ban went into effect after Dobbs and providers stopped performing abortions in WI; Dem WI AG filed a lawsuit to block the ban’s enforcement days after Dobbs, argued at WI Supreme Court in May 2023 |
| Alaska | Legal | Yes |  | In 1997, AK Supreme Court held that abortion rights are fundamental and are encompassed in the state’s recognized right to privacy |
| California | Legal | Yes |  | Abortion remained legal in CA and in 2022 midterms, CA passed Prop. 1 to explicitly protect the right to abortion |
| Colorado | Legal | Yes |  | Abortion legal at all stages of pregnancy; July 6, 2022 EO by Gov. Polis specifying no legal liability for providing or obtaining abortion care |
| Connecticut | Legal | Yes |  | Abortion codified in state law since 1990; right to abortion protected until 24 weeks, after which available if health/life is in danger |
| Delaware | Legal | Yes |  | Abortion legal until viability (law passed in 2017), legal after viability if health/life at risk |
| District of Columbia | Legal | Yes |  | Abortion legal at any stage of pregnancy (DC Council passed law in 2020 to protect reproductive decisionmaking in the District, reaffirmed including wrt abortion in 2022 |
| Hawaii | Legal | Yes |  | Abortion made legal in 1970 – legal until viability, allowed after bortion if health/life at risk |
| Illinois | Legal | Yes |  | Gov. Pritzker signed into law the Reproductive Health Act in 2019 to safeguard abortion; abortion legal pre-viability, allowed post-viability if threat to health or life of woman. Determine legal because abortion was legal for |
| Indiana | Legal |  | Yes | IN passed on 9/15/22 SB1 to ban all abortions except with fetal anomaly, rape/incest, or to save woman’s life, but law blocked on 9/22/22, restoring abortion access up to week 22 except to save woman’s life; judge ruled in 6/23 that abortion ban doesn’t violate state constitution. Included as legal because abortion was legal for 98.3% of the study period, and exceptions for fetal anomaly remained during 1-week ban |
| Iowa | Legal | Yes |  | 2018 IA Supreme Court affirmed a “fundamental right” to abortion but in 2022 it reversed that ruling; IA district court declined in 12/22 Gov. Reynolds’ effort to revive the heartbeat law; in 6/23, IA Supreme Court declined to revive heartbeat law, leaving abortion legal in IA up until 22 weeks |
| Kansas | Legal | Yes |  | In August 2022, voters rejected a proposed constitutional amendment that would have said there’s no right to abortion; KS gov has vetoed three anti-abortion laws |
| Maine | Legal | Yes |  | Abortion legal until viability, after viability permitted if health/life in danger |
| Maryland | Legal | Yes |  | Abortion legal until viability, after viability permitted if health/life in danger, MD Gov signed into law a bill to enshrine abortion rights in May 2023 |
| Massachusetts | Legal | Yes |  | Abortion legal until 24^th^ week, with exceptions allowing abortion later in pregnancy, in 7/22 Gov. Baker signed an abortion bill to protect abortion in MA |
| Michigan | Legal | Yes |  | Abortion legal until viability. In 4/23, Gov. Witmer signed legislation repealing 1931 abortion ban |
| Minnesota | Legal | Yes |  | Abortion protected by MN Constitution; abortion legal until viability, afterwards only to protect life/health |
| Montana | Legal | Yes |  | In 8/22, Montana Supreme Court blocked MT from enforcing a 2021 abortion ban at 20 weeks; in May 2023 Gov. Gianforte signed into law specifying that abortion no longer protected until viability under right to privacy statute |
| Nebraska | Legal | Yes |  | Abortion permitted up to 20 weeks, after that only allowed for life/health of woman; in May 2023 passed a 12-week abortion ban |
| Nevada | Legal | Yes |  | Abortion permitted up to 24 weeks, after that only allowed for life/health of woman |
| New Hampshire | Legal | Yes |  | Abortion permitted up to 24 weeks, after that only allowed for life/health of woman or fetal anomaly |
| New Jersey | Legal | Yes |  | New Jersey Supreme Court has upheld abortion as a constitutional right in NJ for 40 years |
| New Mexico | Legal | Yes |  | No abortion restrictions, abortion legal through all stages of pregnancy |
| New York | Legal | Yes |  | Abortion legal under NY state law since 1970, abortion legal up to 24 weeks or later if health/life at risk or if pregnancy will not survive |
| Oregon | Legal | Yes |  | No abortion restrictions, abortion legal through all stages of pregnancy; abortion codified into state law in 2017 |
| Pennsylvania | Legal | Yes |  | Abortion legal up to 24 weeks, after that only allowed for life/health of woman |
| Rhode Island | Legal | Yes |  | Abortion protected until viability unless health is endangered, abortion right codified in 2019 with passage of Reproductive Privacy Act |
| Vermont | Legal | Yes |  | Abortion protected throughout pregnancy; VT passed in 2019 a law recognizing a fundamental right to abortion |
| Virginia | Legal | Yes |  | Abortion protected through first and second trimesters, allowed in third trimester if woman’s health/life endangered |
| Washington | Legal | Yes |  | Abortion legal in WA since 1970, abortion legal until viability and then only to protect woman’s health/life |
| Wyoming | Legal | Yes |  | Abortion permitted at/after viability only if woman’s health/life endangered; WY banned all abortion except in rape/incest but judge halted the law the day after it went into effect in 3/23; judge prevented WY law banning abortion pills from going into effect in 6/23 |
| Arizona | Middle | Yes | No | Right after Dobbs, abortion providers stopped offering abortions, then resumed abortion care; on 9/23/22 a judge reinstated 19^th^ century total abortion ban in AZ, then on 10/7 a judge halted enforcement of the 1864 anti-abortion law pending litigation, leaving abortion legal up to 15 weeks of pregnancy (they had previously been permitted up to viability) |
| Florida | Middle | Yes | No exception in 15-week ban, narrow exception in heartbeat law | 15-week abortion ban (with exception to safe woman’s life but or if fetal abnormality but no exception for rape/incest) went into effect 7/5/22 after a brief legal challenge accompanied by TRO; 6-week ban signed into law in 4/23 |
| Idaho | Ban | Yes | Yes | Effective 8/25/22, abortion banned by trigger law except when necessary to safe the woman’s life or in cases of rape/incest; on 1/5/23 the ID Supreme Court ruled the ID Constitution doesn’t confer right to abortion; in 4/23 ID became most restrictive state, restricting out-of-state travel for abortion |
| Kentucky | Ban | Yes | No | Near-total abortion ban (unless to save woman’s life, no rape/incest exception) went into effect right after Dobbs (trigger law), in July 2022 a district judge halted enforcement of the law, court of appeals allowed its enforcement in early August 2022, and in February 2023 the near-total abortion ban was allowed by KY Supreme Court to remain in effect |
| North Carolina | Middle | Yes | No | 20-week abortion ban (no rape/incest exception) allowed to go into effect in 8/22, 12-week ban went into effect 7/1/23 |
| Ohio | Middle | Yes | No | Abortions allowed through 20 weeks gestation (no rape/incest exception, only exception for medical emergency) because heartbeat law that went into effect right after Dobbs was temporarily blocked in 9/22 and 10/22, blocked again in 12/22 by court of appeals |
| South Carolina | Middle | Yes | No | Abortion banned after 20 weeks postfertilization, heartbeat bill passed in 5/23 but blocked by state court pending state Supreme Court review |
| Utah | Middle | Yes | No | Abortion legal up to 18 weeks except to safe life/health of woman or fetus will not survive (no rape/incest exception), in 5/23 judge blocked law that would have forced all abortion clinics to close |
| West Virginia | Middle |  | Only up to 8 weeks | In 7/22 a judge blocked enforcement of 150-year-old abortion ban, allowing abortions to resume because of conflict with 2015 law allowing abortion up until 20 weeks postfertilization except to save woman’s life (but only one clinic left); WV Gov signed into law in 9/22 a near-total abortion ban with rape exception up to 8 weeks and medical exception |

Notes: Some grouping decisions differ from known public resources; for example, one resource currently lists Indiana as restricted because a law restricting abortions went into effect in August of 2023; we included it in the legal state grouping because abortion was technically legal there for >98.3% of the excess mortality study period.

**Figure S1. Fitted deaths per 100,000 births for each state group by Joinpoint analysis**

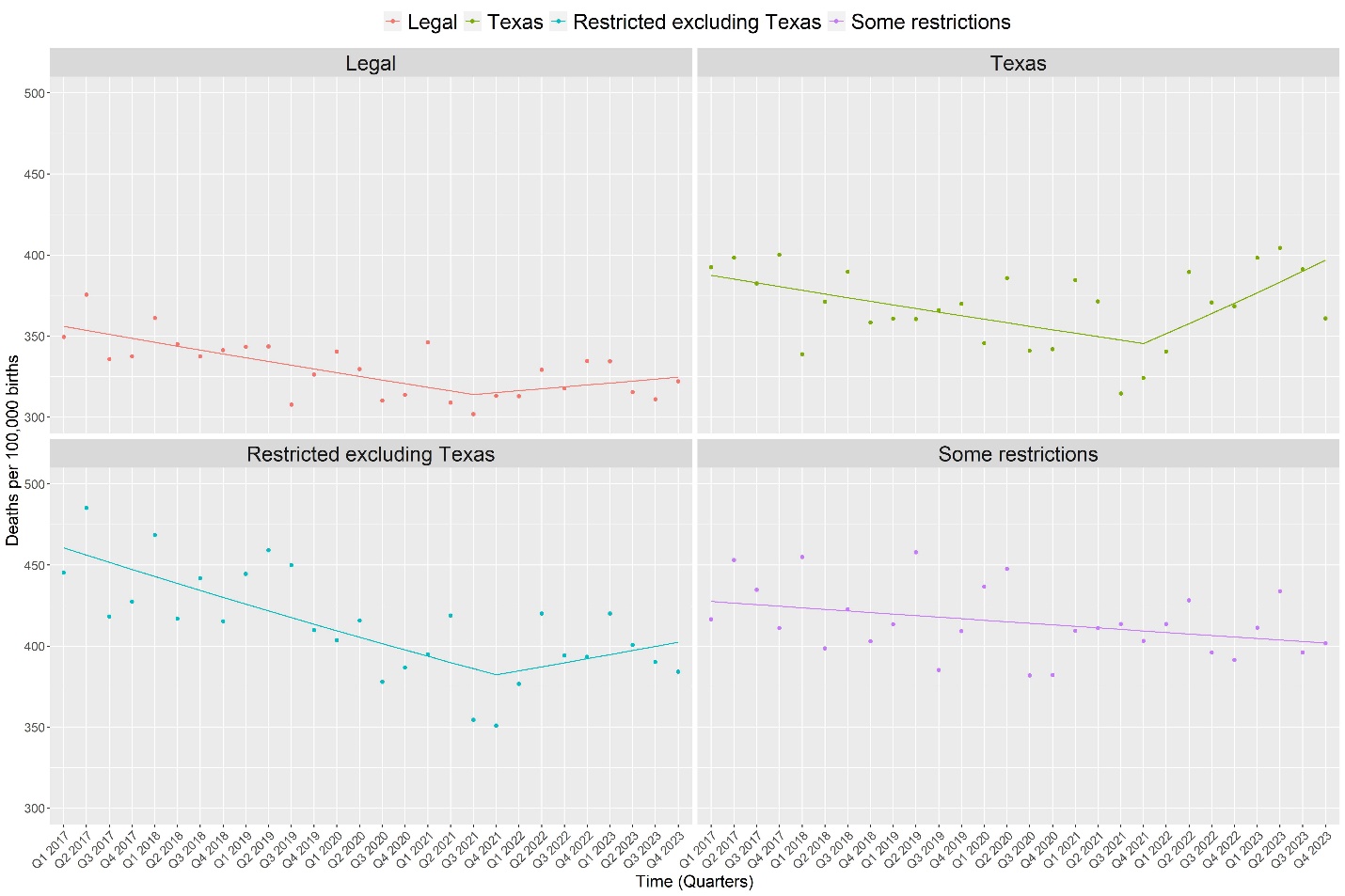

Observed (dots) and fitted deaths (lines) per 100,000 births in Legal states, Texas, Restricted (excluding Texas) states, and Some restrictions states. Quarterly data January 1, 2017-September 30, 2023) were batched. The model spontaneously identified joinpoints in Legal states, Texas, and Restricted (excluding Texas) states.

**Table S2. Statistical testing for joinpoint analysis.**

| **State** | **joinpoint** | **slope before joinpoint** | **slope after joinpoint** | **Nominal p-value** | **adjusted p-value** |
| --- | --- | --- | --- | --- | --- |
| Legal | Q3, 2021 | -2.335 | 1.189 | 0.003 | 0.012 |
| Texas | Q4, 2021 | -2.214 | 6.428 | 0.002 | 0.008 |
| Restricted excluding Texas | Q4, 2021 | -4.122 | 2.509 | 0.007 | 0.028 |
| Some restrictions | none | overall slope = -0.946 | | - | - |

Note 1: Adjusted p-value by Bonferroni correction was calculated to control the Family-wise error rate. Adjusted p values <0.05 indicate joinpoint statistical significance.

Note 2: As results were derived from logistic regression rather than a linear model, slopes are not constant over time. The slopes reported in the above table represent the average slope during specific periods.

**Table S3. sARIMA-model inputs and internal synthetic control derived excess mortality results.**

| **State** | **Births** | **Expected deaths (95% CI)** | **Observed Deaths** | **Observed Deaths per 10,000 Births** | **Excess deaths, no. (95% CI)** | **Ratio of observed to expected deaths (95% CI)** | **Excess Deaths per 10,000 Births** |
| --- | --- | --- | --- | --- | --- | --- | --- |
| Legal | 3,819,879 | 12,053 (11,014 - 13,092) | 12,303 | 32.2 | 250 (-789 - 1,289) | 1.02 (0.94 - 1.12) | 0.7 (-2.1 - 3.4) |
| Restricted excluding Texas | 1,177,522 | 4,285 (3,596 - 4,974) | 4,677 | 39.7 | 392 (-297 - 1,081) | 1.09 (0.94 - 1.30) | 3.3 (-2.5 - 9.2) |
| Some restrictions | 1,488,040 | 6,074 (5,639 - 6,509) | 6,080 | 40.9 | 6 (-429 - 441) | 1.00 (0.93 - 1.08) | 0 (-2.9 - 3) |
| Texas | 777,615 | 2,649 (2,379 - 2,920) | 2,938 | 37.8 | 289 (18 - 559) | 1.11 (1.01 - 1.24) | 3.7 (0.2 - 7.2) |

**Figure S2. Share of newborns not alive at time of birth certificate by race/ethnicity.**
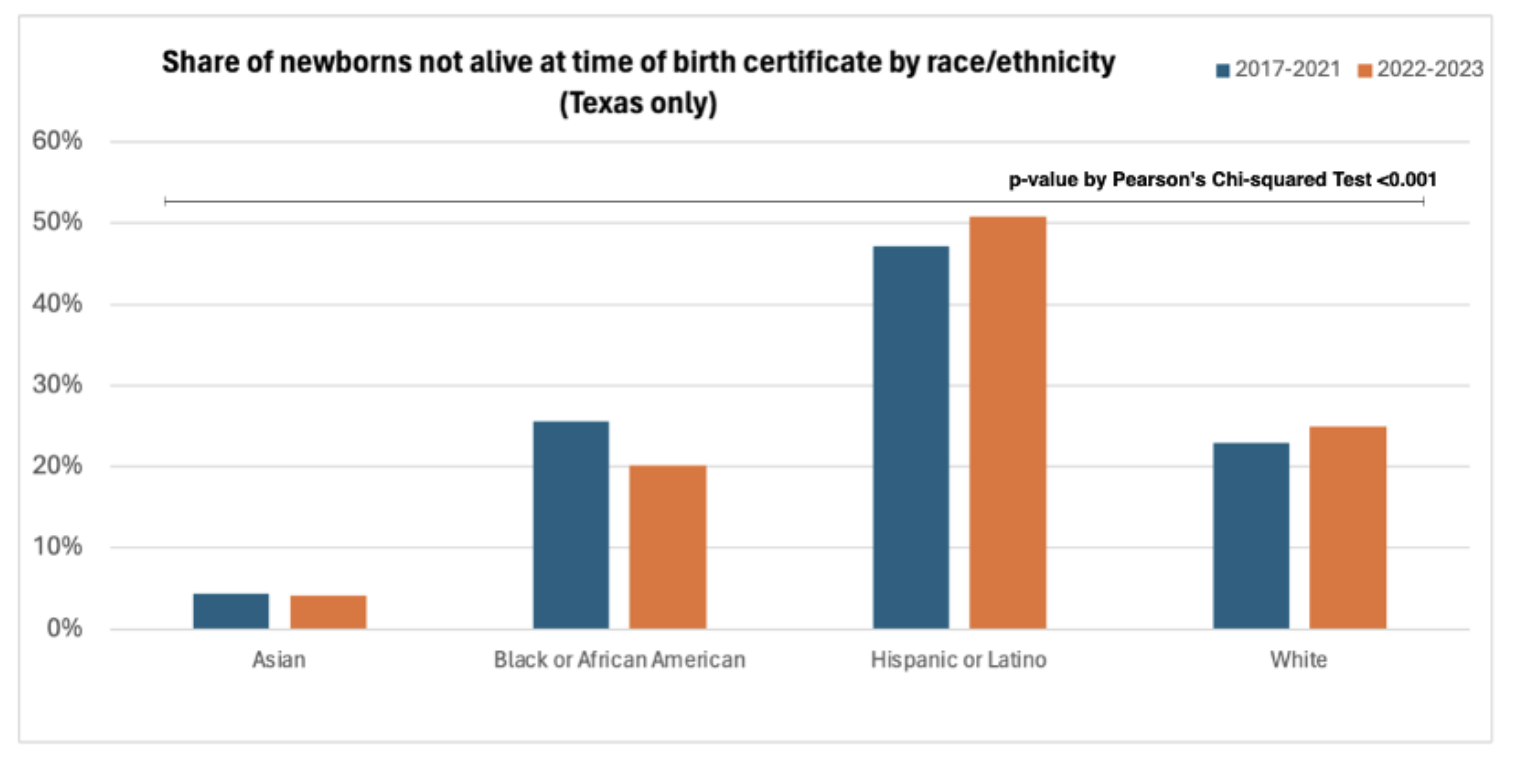

Shares (by race/ethnicity for groups with adequate data) of newborns reported ‘not alive’ at the time of completion of the birth certificate. Data from 2017-2021 (blue bars) and 2022-2023 (orange bars) are shown. All Pearson’s Chi-Square test differences were significant. *Note: While 2023 data are incomplete, the use of births as denominators were used, minimizing lag eff*ects.
